# SEVERITY OF LUNG OBSTRUCTION AND OLDER AGE, BUT NOT PHYSICAL ACTIVITY, PREDICT LOCOMOTOR MUSCLE OXIDATIVE IMPAIRMENT IN COPD

**DOI:** 10.1101/2025.01.28.25321301

**Authors:** Alessandra Adami, Fenghai Duan, Robert A. Calmelat, Zeyu Chen, Richard Casaburi, Harry B. Rossiter

## Abstract

**Background:** Low muscle oxidative capacity is an extrapulmonary manifestation of chronic obstructive pulmonary disease (COPD) with unclear etiology. We sought to determine clinical and behavioral features associated with muscle oxidative capacity in smokers with and without COPD and never smoker individuals.

**Methods:** 243 adults enrolled in the *Muscle Health Study*, an observational study ancillary to COPDGene. G*astrocnemius* oxidative capacity was measured by near-infrared spectroscopy from muscle oxygen uptake recovery rate constant (*k*). Physical activity by accelerometry (vector magnitude units, VMU/min). Pulmonary assessments included spirometry (FEV_1_%predicted), diffusing capacity (DL_CO_), and quantitative chest computed tomography (CT). Eighty-seven variables related to COPD features were considered. Variables selected by univariate analysis of log-transformed *k* with p≤0.20, and filtered by machine learning, were entered into multivariable linear regression to determine association with *k*.

**Results:** 241(99%) participants were allocated to analysis. FEV_1_%predicted, DL_CO_, CT, pack-years, age and VMU/min were among 24 variables selected by univariate analysis. After machine learning filtering on 161(66%) cases with complete data, 11 variables were included in multivariable analysis. Only FEV_1_%predicted, age and race were significantly associated with *k* (R^2^=0.26). Model coefficients equate a 10% lower FEV_1_%predicted to a 4.4% lower *k*, or 10-years of aging to a 9.7% lower *k*. In 118 cases with CT available, FEV_1_%predicted and age remained associated with *k* (R^2^=0.24). Physical activity was not retained in any model.

**Conclusions:** Locomotor muscle oxidative capacity was positively associated with FEV_1_%predicted and negatively associated with age. Physical activity or radiographic COPD manifestations were not significantly associated with muscle oxidative impairment.

## INTRODUCTION

In people with chronic obstructive pulmonary disease (COPD), impaired exercise capacity, skeletal muscle dysfunction and physical inactivity are strong predictors of increased healthcare utilization, poor quality-of-life and morbidity[1-3]. Extra-pulmonary COPD manifestations, e.g., low bone density, muscle atrophy and weakness, low muscle oxidative capacity and capillarity, are also associated with mortality[2, 4]. Locomotor muscle structural changes (typically assessed in quadriceps) include atrophy, an increase in reactive oxygen species production and glycolytic type II fibers, loss of fatigue-resistant type I fibers, and ∼10-50% loss of oxidative capacity[2, 5, 6]. These adaptations may underlie the strong association between exercise intolerance and mortality in COPD[7, 8]. However, the etiology of the muscular defect in oxidative function is still not well understood[3].

A large number of factors are proposed to contribute to loss of muscle oxidative capacity in COPD, from a normal deconditioning response to low physical activity (i.e., non-pathologic) to systemic factors (e.g., inflammation) related to smoking and/or pulmonary obstruction severity[7, 9]. Muscle disuse leads to many of the changes observed in COPD including muscle atrophy, reduced oxidative capacity and vasoreactivity[10]. It remains unclear whether behavioral or biologic processes contribute to loss of the oxidative phenotype in COPD[3].

Skeletal muscle oxidative capacity may be estimated noninvasively by near-infrared spectroscopy (NIRS)[11]. This relies on a property of activated muscle mitochondria, that the recovery rate constant (*k*, min^-1^) of muscle oxygen consumption following contractions is directly proportional to muscle oxidative capacity. This concept has been validated in single isolated muscle fibers and in comparison to muscle biopsy and ^31^P magnetic resonance spectroscopy[12]. NIRS-based methods directly assess muscle cellular capacity for oxidation under occlusion and its measurement is therefore isolated from pathological circulatory or pulmonary derangements observed in COPD[11].

We sought to determine clinical and behavioral features associated with muscle oxidative capacity in smokers with COPD across the range of disease severity, smokers without COPD, and never smoker individuals. We hypothesized that low physical activity and older age would associate with low muscle oxidative capacity.

## METHODS

### Participants

We enrolled 243 adults, of whom 218 were current or former smokers with ≥10 pack-years smoking history. Recognizing that smokers without COPD do not represent a “healthy” condition, 76 smokers with normal spirometry were used as a COPD comparator group, in attempt to control for influence of smoking history. Twenty-seven participants were never smokers (lifetime <100 cigarettes, <52 cigars, or <12oz. pipe tobacco smoked).

Smokers with normal spirometry and never smoker individuals had post-bronchodilator forced expiratory volume in 1 second (FEV_1_) and forced vital capacity (FVC) ≥0.70 and FEV_1_ ≥80%predicted. Preserved ratio impaired spirometry (PRISm) individuals had FEV_1_/FVC ≥0.70 but FEV_1_ <80%predicted. Participants with COPD had FEV_1_/FVC <0.70. All subjects were in stable state, with no exacerbation within 4-weeks of enrollment. Exclusion criteria included participation in pulmonary rehabilitation within 18-months, pregnancy, or significant disease other than COPD that may: (i) put the person at risk by participation; (ii) influence study results, such as ischemic heart disease, musculoskeletal or renal disease; or (iii) limit the individual’s ability to comply with study protocol.

Participants were informed about study procedures and risks before giving written informed consent. The study was approved by the Institutional Review Board of The Lundquist Institute at Harbor-UCLA Medical Center (20403-01).

### Study design

The *Muscle Health Study*, a single-center observational study and ancillary study of COPDGene[13] (NCT00608764), took place between 2014 and 2016 at the Respiratory Research Center at the Lundquist Institute, Torrance, CA, USA. We enrolled 224 individuals who were also COPDGene participants and 19 were recruited from Harbor-UCLA Medical Center pulmonary outpatient clinic (Figure 1).

**Figure 1.**
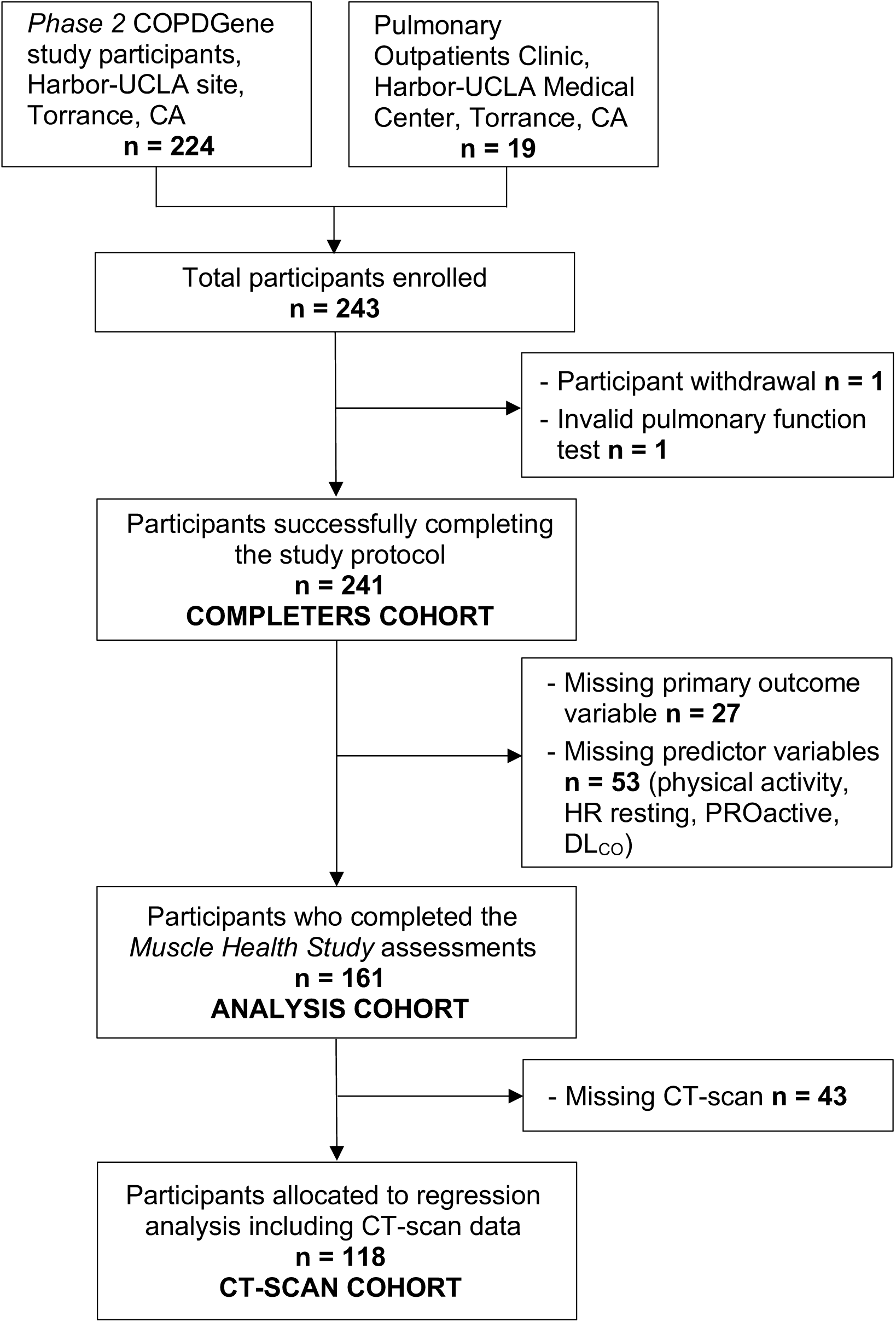
Study flow diagram. Study enrollment and allocation of participants to final analysis based on completion of the muscle test to determine skeletal muscle oxidative capacity (*study primary outcome*) and physical activity monitoring.

Participants visited the laboratory once and completed, among other assessments: spirometry; lung diffusing capacity for carbon monoxide (DL_CO_); inspiratory and expiratory chest computed tomography (CT) imaging; six-minute walking distance (6WMD); questionnaires related to medical and smoking history, occupational and educational background, symptoms (COPD Assessment Test, CAT; modified Medical Research Council Dyspnea scale, mMRC), health-related quality-of-life (St. George’s Respiratory Questionnaire, SGRQ; 36-Item Short Form Health Survey, SF-36), and anxiety and depression (Hospital Anxiety and Depression Scale, HADS); NIRS-based muscle oxidative capacity; resting pulse oximetry monitoring; free-living physical activity (PA) monitoring; and the Daily-PROactive Physical Activity in COPD (D-PPAC) instrument[14].

Detailed description of the methods is available in the Supplementary material.

### Pulmonary function and imaging

Spirometry was performed in accordance with American Thoracic Society (ATS) guidelines[15]. DL_CO_, measured in accordance with European Respiratory Society/ATS standards[16], was adjusted for haemoglobin and altitude, and relative (%predicted) values were calculated using Global Lung Initiative reference equations[17].

Chest CT scans were performed in the supine position during a single full inspiration (200 mAs) and at end-tidal exhalation (50 mAs)[13]. Airway wall thickening, %emphysema, %gas trapping, and square root of wall area of a 10-mm perimeter airway (Pi10) [18, 19] were determined.

### NIIRS muscle oxidative capacity

The medial *gastrocnemius* muscle was assessed using continuous-wave, spatially-resolved spectroscopy NIRS (PortaMon, Artinis BV, NL). NIRS measures relative concentration of deoxygenated (HHb+Mb) and oxygenated (HbO_2_+MbO_2_) hemoglobin and myoglobin within the muscle up to a depth of ∼1.5 cm under the probe. From these measurements, relative changes in total hemoglobin and myoglobin (THb = HHb+Mb+HbO_2_+MbO_2_) were calculated. Tissue saturation index (TSI, an index of tissue oxygen saturation) was calculated using spatially-resolved spectroscopy.

With the participant supine on an exam bed, the belly of the calf muscle was identified. Palpation during a series of brief isometric muscle contractions was used to optimize placement of the plastic-wrapped NIRS probe longitudinally over a highly activated region of the medial *gastrocnemius.* The NIRS probe was then secured in position using an elastic bandage. A rapid-inflation pressure-cuff (SC12D, Hokanson, USA) was placed on the proximal thigh on the same limb as the NIRS probe and attached to an electronically-controlled rapid cuff-inflator (E20, Hokanson, USA). The participant was asked to relax and minimize limb movement, except when instructed. The participant was then familiarized with rapid cuff-inflation and to the brief muscle contractions required in the protocol. Arterial occlusion pressure was determined from a tolerated cuff-pressure within the range of 153-300 mmHg (mean: 221±24 mmHg) that resulted in a rise in HHb+Mb, a fall in HbO_2_+MbO_2_ and stable THb signal over 15-20 s. Repeated dynamic muscle contractions were made at ∼1 Hz by plantar-flexion/relaxation against a light resistance (hereafter referred to as muscle contractions).

Initially, the participant laid at rest for 2-3 min to establish a stable baseline TSI. After this, ∼10-15 s of light muscle contractions were performed to increase mV̇O_2_, following which blood flow was occluded until a stable minimum TSI was reached or for 5 min (whichever occurred first). Cuff pressure was then released and the subsequent reactive reoxygenation monitored until resting baseline was reestablished (typically ∼3 min[11]). This was used to determine an individualized muscle saturation physiological range (see Figure 1 in[11]). Finally, two muscle oxidative capacity assessments were performed, each consisted of: 1) ∼10-15 s muscle contractions to increase mV̇O_2_ and desaturate the muscle to ∼50% of the physiological range[11]; 2) a series of intermittent arterial occlusions (5 occlusions for 5 s, 10 for 10 s, each separated by 5-20 s recovery). Each of the assessments lasted ∼6 min and were separated by ∼2 min of rest.

For each intermittent arterial occlusion during oxidative capacity tests, the linear rate of decline in TSI (desaturation in %.s^-1^) was determined and a point value for relative mV̇O_2_ reported. Because the rate of tissue deoxygenation during arterial occlusion is inversely proportional to mV̇O_2_, its value is reported as positive (%.s^-1^; see Figure 2 in[11]). The primary study outcome was the exponential mV̇O_2_ recovery rate constant (*k*, min^-1^), which was estimated using non-linear least-squares regression[11] (OriginPro v8.6, OriginLab Co., Northampton, USA). This protocol has good test–retest reliability in smokers with and without COPD (ICC ≥0.88[11, 12]).

**Figure 2.**
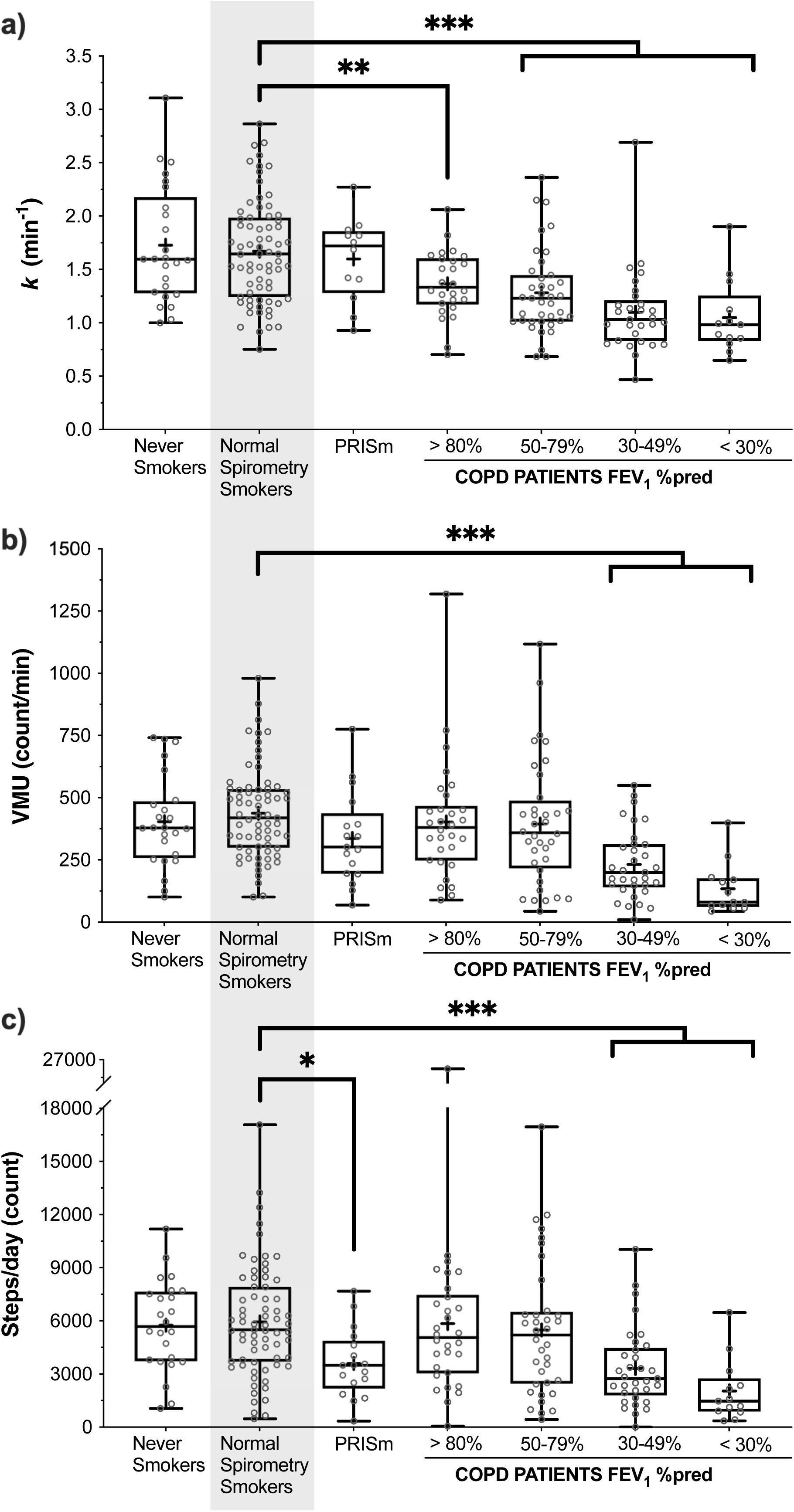
Comparison of (**a**) the rate constant of muscle oxygen consumption recovery (*k;* proportional to muscle oxidative capacity; n=214), (**b**) physical activity (mean vector magnitude units per minute; n=222), and (**c**) number of steps per day (n=222), among smokers with normal spirometry (reference group; Controls), never smokers, and individuals with PRISm and COPD. *k*, rate constant of muscle oxygen consumption recovery; VMU, vector magnitude units; PRISm, preserved ratio impaired spirometry; COPD, chronic obstructive pulmonary disease; FEV_1_ %pred, forced expiratory volume in 1 second as a percentage of the predicted value. *p<0.05; ** p<0.01; *** p<0.001.

### Pulse oximetry

Arterial oxygen saturation was estimated at rest prior to the NIRS test using fingertip pulse oximetry (SpO_2_) (Rad-5 MasimoSET®, Masimo Co., USA).

### Free-living daily physical activity

Participants underwent 7 days of PA monitoring using triaxial accelerometry (Dynaport MoveMonitor, McRoberts BV, NL). Each participant was instructed in the correct positioning of the monitor in small of the back, and to adjust the elastic waistband to ensure the device was in contact with the body and comfortable. Participants were asked to wear the PA monitor for as long as possible during each 24-hour period, and to remove it only for bathing or swimming. The PA monitor was worn from the end of the study visit until 7 full days had elapsed. Data were processed using the manufacturer’s protocols.

Daily PA measurements were accepted as valid if they met the criteria that the monitor was worn for at least 8 hours/day during waking hours[14] and for at least 4 days/week (not necessarily consecutive, without distinction between weekdays and weekends). Compliance with these conditions was 93%. PA is reported as the mean number of steps per day (total accumulated during each daytime period), and as the mean count of vector magnitude units per minute (VMU/min) during “daytime” hours between 8 AM and 11 PM[20]. VMU is the vectorial sum of movements in the three orthogonal planes (sagittal, frontal, transversal) measured over a one-minute epoch. The device includes an accelerometer sensor sensitive to these orthogonal axes. The acceleration is measured while the device is worn, the signal is converted to a digital representation and processed to obtain an “activity count”, i.e. the VMU. Sample frequency of the triaxial accelerometer is automatically set to 100Hz.

Subjective PA experience was determined using the D-PPAC, on the same days the accelerometer was worn[14]. D-PPAC results are presented as Rassh scores (0 (worst)–100 (best)) for subdomains of PA amount and difficulty, and their sum total [14].

### Statistical Analyses

Data are presented as mean(SD) for continuous variables, counts and percentage for discrete variables. One-way ANOVA with Dunnett’s *post hoc* determined differences from smokers with normal spirometry (here termed ‘Controls’). To determine effect of chronic and acute smoking (defined as having smoked at least one cigarette in the 2-hours prior to the NIRS test) in normal spirometry participants, 1-way ANOVA and unpaired t-test were performed with Bonferroni correction for multiple comparisons.

To determine association of predictor variables with the recovery rate constant of muscle oxygen consumption (*k*; proportional to muscle oxidative capacity), a univariate linear regression analysis was performed on the completers cohort (n=241) with *k* log transformed to satisfy the homoscedasticity regression assumption. Collinear variables, variables with more than 30% missing data, and comorbidities with reported frequency of zero within the completers cohort (cancer of the kidney, cancer of the throat and mouth, connective tissue disorder, ovarian cancer, pancreatic cancer), were excluded from subsequent analysis.

Then, variables with p≤0.20 in univariate linear regression were further filtered using machine learning-based feature selection methods (LASSO, Random Forest, XGBoost). These methods were selected to enhance the reliability of the identified variables and to account for more complex dependencies and interactions among predictors’ effects on the response variable. A total of 13 predictors were identified to be included in the multivariable linear regression model. To address multicollinearity, two variables with high variance inflation factor (VIF) values were excluded. The remaining 11 top-ranked variables were entered into the final multivariable linear regression to identify predictors of *k*. Due to a relatively large number of participants (n=43) who opted out of CT imaging, a subset analysis was performed on those with a complete dataset that included CT imaging (Figure 1). Additional details are provided in the Supplementary material. The significance level (α) was set at 0.05.

## RESULTS

### Participants

Of 243 enrolled, one withdrew, one was excluded for invalid pulmonary function test, and 241(99.2%) successfully completed study assessments (‘completers cohort’; Figure 1). Table 1 reports characteristics of the completers and includes comparison across sub-groups based on smoking history and obstruction severity.

**Table 1.** Personal characteristics of participants completing the study protocol (completers cohort; n=241). Comparison is among smokers with normal spirometry (reference group; ‘Controls’), never smokers, preserved ratio impaired spirometry, and patients with COPD (subdivided by disease severity stage).

|  | COHORT | Never Smokers | Normal Spirometry Smokers (Controls) | PRISm | COPD FEV<sub>1</sub> %predicted |  |  |  |
| --- | --- | --- | --- | --- | --- | --- | --- | --- |
|  |  |  |  |  | > 80 | 50-79 | 30-49 | < 30 |
| N. | 241 | 26 | 76 | 21 | 30 | 40 | 35 | 13 |
| **Personal characteristics** |  |  |  |  |  |  |  |  |
| Age (yrs) | 63.9 (9.8) | 59.4 (10.3) | 60.0 (8.7) | 64.0 (11.2) | 70.1 (9.6) * | 65.7 (8.1) * | 68.0 (8.0) * | 65.2 (10.1) |
| Height (cm) | 168.8 (9.8) | 167.6 (8.3) | 160.1 (9.7) | 168.3 (9.3) | 169.8 (10.5) | 171.5 (9.6) | 167.1 (11.2) | 170.2 (9.5) |
| Weight (kg) | 80.8 (18.6) | 79.6 (15.3) | 82.0 (20.8) | 90.8 (18.6) | 76.9 (15.3) | 81.6 (16.9) | 77.5 (19.4) | 75.2 (17.8) |
| BMI (kg.m<sup>-2</sup>) | 28.2 (5.9) | 28.3 (4.4) | 28.6 (6.7) | 31.1 (5.7) | 26.6 (4.1) | 27.8 (5.3) | 27.7 (6.3) | 26.3 (7.2) |
| Sex (F/M) | 128/113 | 15/11 | 42/34 | 14/7 | 17/13 | 18/22 | 18/17 | 4/9 |
| Race (AA/As/Oth/W) | 110/1/1/129 | 13/0/0/13 | 50/0/0/26 | 10/0/0/11 | 8/0/0/22 | 13/0/0/27 | 12/1/1/21 | 4/0/0/9 |
| Ethnicity (H/Not-H) | 3/238 | 0/26 | 0/76 | 0/21 | 0/30 | 0/40 | 2/33 | 1/12 |
| **Pulmonary function, Medical and smoking history** |  |  |  |  |  |  |  |  |
| FVC (l) | 3.1 (0.9) | 3.5 (0.8) | 3.3 (0.8) | 2.3 (0.5) * | 3.7 (0.8) | 3.1 (0.6) | 2.5 (0.8) * | 2.2 (0.6) * |
| FEV<sub>1</sub> (l) | 2.1 (0.9) | 2.9 (0.7) | 2.7 (0.6) | 1.8 (0.4) * | 2.4 (0.6) | 1.9 (0.5) * | 1.0 (0.3) * | 0.6 (0.2) * |
| FEV<sub>1</sub>/FVC | 0.66 (0.17) | 0.81 (0.06) | 0.80 (0.05) | 0.76 (0.05) | 0.64 (0.06) * | 0.60 (0.06) * | 0.43 (0.10) * | 0.29 (0.09) * |
| FEV<sub>1</sub> % pred | 78.8 (28.5) | 106.5 (13.8) | 101.0 (12.5) | 69.1 (10.9) * | 92.9 (11.1) | 66.2 (10.0) * | 40.0 (5.6) * | 21.0 (4.3) * |
| DL<sub>CO</sub> % pred | 70.4 (22.6) | 88.8 (10.8) | 80.1 (17.0) | 65.8 (13.2) * | 75.5 (19.5) | 66.2 (24.1) * | 44.3 (12.6) * | 37.2 (14.4) * |
| Hemoglobin (g/dL) | 13.7 (1.4) | 13.6 (1.2) | 13.7 (1.3) | 13.5 (1.4) | 13.7 (1.1) | 13.9 (1.5) | 13.7 (1.7) | 14.2 (1.7) |
| Resting SpO<sub>2</sub> (%) | 97.5 (2.3) | 98.7 (0.5) | 98.4 (1.1) | 97.1 (3.0) | 97.6 (2.2) | 97.4 (1.3) | 96.3 (2.3) * | 94.5 (5.5) * |
| Supplementary O<sub>2</sub> (N (%)) | 26 (10.8) | 0 (0) | 0 (0) | 1 (5) | 1 (3) | 1 (3) | 14 (40) | 12 (92) |
| Smoking duration (yrs) | 37.5 (10.1) | - | 34.4 (9.3) | 39.0 (13.1) | 38.9 (10.6) | 39.8 (9.0) * | 41.5 (9.7) * | 33.9 (8.2) |
| Smoking history (pack-yrs) | 44.4 (22.7) | - | 39.4 (19.8) | 43.6 (25.7) | 39.8 (18.4) | 52.7 (25.2) * | 52.9 (20.9) * | 41.9 (28.6) |
| Smoking status (NS/CS/FS) | 27/82/132 | 26/0/0 | 0/42/34 | 0/10/11 | 0/9/20 | 0/13/27 | 0/8/27 | 0/0/13 |
| **Functional performance and Activity** |  |  |  |  |  |  |  |  |
| BODE | 1.7 (2.2) | 0.4 (0.8) | 0.9 (1.2) | 1.6 (1.8) | 0.5 (1.1) | 1.7 (1.5) * | 5.2 (1.5) * | 6.9 (2.0) * |
| 6MWD (m) | 396 (107) | 496 (95) * | 400 (81) | 380 (97) | 445 (107) | 401 (70) | 324 (109) * | 249 (105) * |
| Recent exercise (N (%)) | 135 (56.3) | 20 (76.9%) | 41 (54.0%) | 12 (57.1%) | 18 (60.0%) | 25 (62.5%) | 13 (38.2%) | 6 (46.2%) |
| D-PPAC Total score | 62.3 (12.5) | 72.0 (13.1) | 65.3 (10.1) | 62.3 (8.0) | 65.7 (11.6) | 64.9 (10.5) | 52.5 (9.0) * | 47.1 (14.1) * |
| Amount score | 48.5 (13.6) | 53.7 (16.0) | 51.7 (12.0) | 49.7 (9.0) | 52.8 (10.8) | 49.3 (12.3) | 40.9 (11.2) * | 31.5 (15.5) * |
| Difficulty score | 76.1 (17.5) | 89.9 (13.9) * | 78.2 (16.3) | 74.1 (18.0) | 77.9 (16.8) | 79.9 (17.5) | 63.5 (12.0) * | 62.1 (15.1) * |

Data are mean (SD) if not otherwise specified. BMI, body mass index; F, female; M, Male; AA, African American; As, Asian; Oth, other races; W, White Caucasian; H, Hispanic or Latino; Not-H, Not-Hispanic or Latino; FVC, Forced vital capacity; FEV<sub>1</sub>, forced expiratory volume in 1 second ; DL<sub>co</sub>, Lung diffusing capacity for carbon monoxide adjusted for [hemoglobin] and altitude; SpO<sub>2</sub>, oxygen saturation; NS, never smoker; CS, current smoker; FS, former smoker; BODE, index derived from BMI, airflow obstruction, dyspnea and exercise (6-minute walk distance); 6MWD, 6-minute walking distance (in meters); Recent exercise, defined as having done exercise in the past 3 weeks (Yes/No); D-PPAC, Daily PROActive Physical Activity instrument in COPD (score 0-100). Spirometric variables are postbronchodilator. \* p<0.05 versus normal spirometry smokers (controls)

Of 241 participants, 128(53.1%) were female, 110(45.6%) African American, and 3(1.2%) of Hispanic ethnicity. Participants were 64±10 years old and overweight (BMI 28.2±5.9 kg/m^2^); 82(34.0%) were current smokers, 54.8% former smokers and 10.8% never smokers. 150(62.2%) participants had one or more comorbidities (Table S1), and 26(10.8%) were prescribed supplementary O_2_ therapy. Also, 134(55.6%) reported having participated in exercise (defined as walking or biking for exercise at least twice a week within the previous 3-weeks).

Overall, there were 26 never smokers, 76 Controls, 21 PRISm, and 118 COPD (Table 1). COPD participants were older than Controls, had significantly lower resting SpO_2_ and exercise performance (6MWD), and worse dyspnea symptoms and quality-of-life (BODE, mMRC, CAT, SGRQ total score) (p<0.001; Table S2). Never smokers had significantly greater 6MWD and reported less difficulty with PA (D-PPAC difficulty score) compared to Controls. PRISm reported worse physical functioning perception (SF-36 physical functioning and component scores; p<0.045; Table S2) but not significant differences in symptoms and quality-of-life (p>0.379) despite being worse than Controls.

Eight COPD patients required nasal cannula O_2_ during the study visit (2-4 L.min^-1^; note that this has been found to not influence NIRS-based muscle oxidative capacity assessment[11]).

### Muscle oxidative capacity

Muscle oxidative capacity was reliably assessed in 214(88.8%) participants. Twenty-one were unable to tolerate the pressure for arterial occlusions, while *k* could not be determined from recorded measurements in six tests.

On average, *k* was not different among never smokers (1.73±0.55 min^-1^; n=25), Controls (1.67±0.48 min^-1^; n=70) and PRISm (1.60±0.39 min^-1^; n=12) (Figure 2a). Compared to Controls, *k* was 17.9% lower in mild COPD (1.37±0.31 min^-1^; n=26; p<0.003), and was progressively impaired with increasing COPD severity (moderate, 1.28±0.40 min^-1^, n=39; severe, 1.10±0.40 min^-1^, n=29; very-severe, 1.05±0.35 min^-1^, n=13; each p<0.0001) (Figure 2a).

To address whether impaired *k* in COPD was related to direct effects of smoke exposure rather than COPD *per se*, we investigated the acute effect of smoking on *k* by comparing subjects who had vs. had not smoked within 2 hours and also the chronic effect of smoking (>10 pack-years) in smokers with normal spirometry in comparison to never smokers. We found no difference in *k* (p=0.324) among never smokers (1.73±0.55 min^-1^; n=25), current smokers (1.75±0.52 min^-1^, n=38) or former smokers (1.58±0.43 min^-1^, n=32) with normal spirometry (Figure 3a). Within normal spirometry current smokers (n=70), we found no difference in *k* (p=0.785) between individuals who had smoked within 2 hours (1.70±0.34 min^-1^, n=21) and those who had not (1.74±0.55 min^-1^, n=17) (Figure 3b).

**Figure 3.**
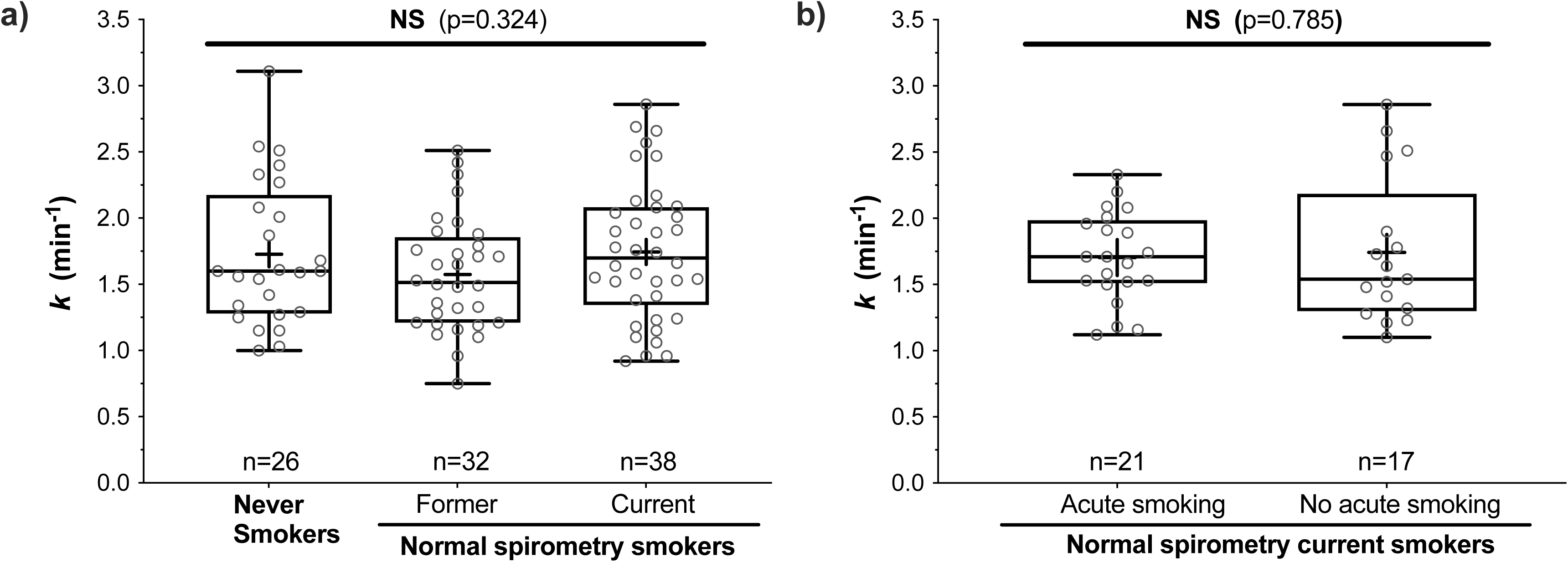
Chronic (**a**) and acute (**b**) effects of smoking on the rate constant of muscle oxygen consumption recovery (*k*; proportional to muscle oxidative capacity) in participants with normal spirometry.

### Free-living daily physical activity and symptoms

A total of 222(92.1%) wore the triaxial accelerometer for at least 8 daytime hours over an average of 5.6±1.4 days. As expected, PA was lowest in severe COPD, who walked ≥44% fewer steps/day (severe, 3328±2241, n=33; very-severe, 2037±1756, n=13; p<0.0001) and moved ≥47% less on average (VMU: severe, 232±139 counts/min, n=33; very-severe, 135±73 counts/min, n=13; p<0.0001) than Controls (5938±3121 steps/day; 438±184 counts/min, n=69) (Figure 2b and 2c). Of interest, steps/day were also lower in PRISm than Controls (3599±1928, n=17; p=0.0022; Figure 2c). Otherwise, PA behaviors in never smokers, mild and moderate COPD did not differ from Controls (Figure 2b and 2c).

Severe COPD patients also reported lower activity amount, difficulty and total score using D-PPAC compared with Controls (p<0.001) (Table 1), indicating greater difficulty in sustaining the limited activity they performed. Similarly, physical functioning and physical component scores of SF-36 were significantly (p<0.001) lower in severe COPD compared with Controls (Table S2).

Moderate and severe COPD reported worse symptoms (mMRC, CAT) and quality-of-life (SGRQ) compared with Controls (p<0.05; Table S2).

### Predictors of muscle oxidative capacity

Univariate and multivariable linear regression analyses identified muscle oxidative capacity predictors. A total of 241 participants were allocated to univariate analysis (‘completers cohort’; Figure 1). Eighty-seven variables were considered in the univariate regression analysis, of which 24 were significantly associated with *k*; the strongest were age, DL_CO_ and spirometric variables (Table 2).

**Table 2.**
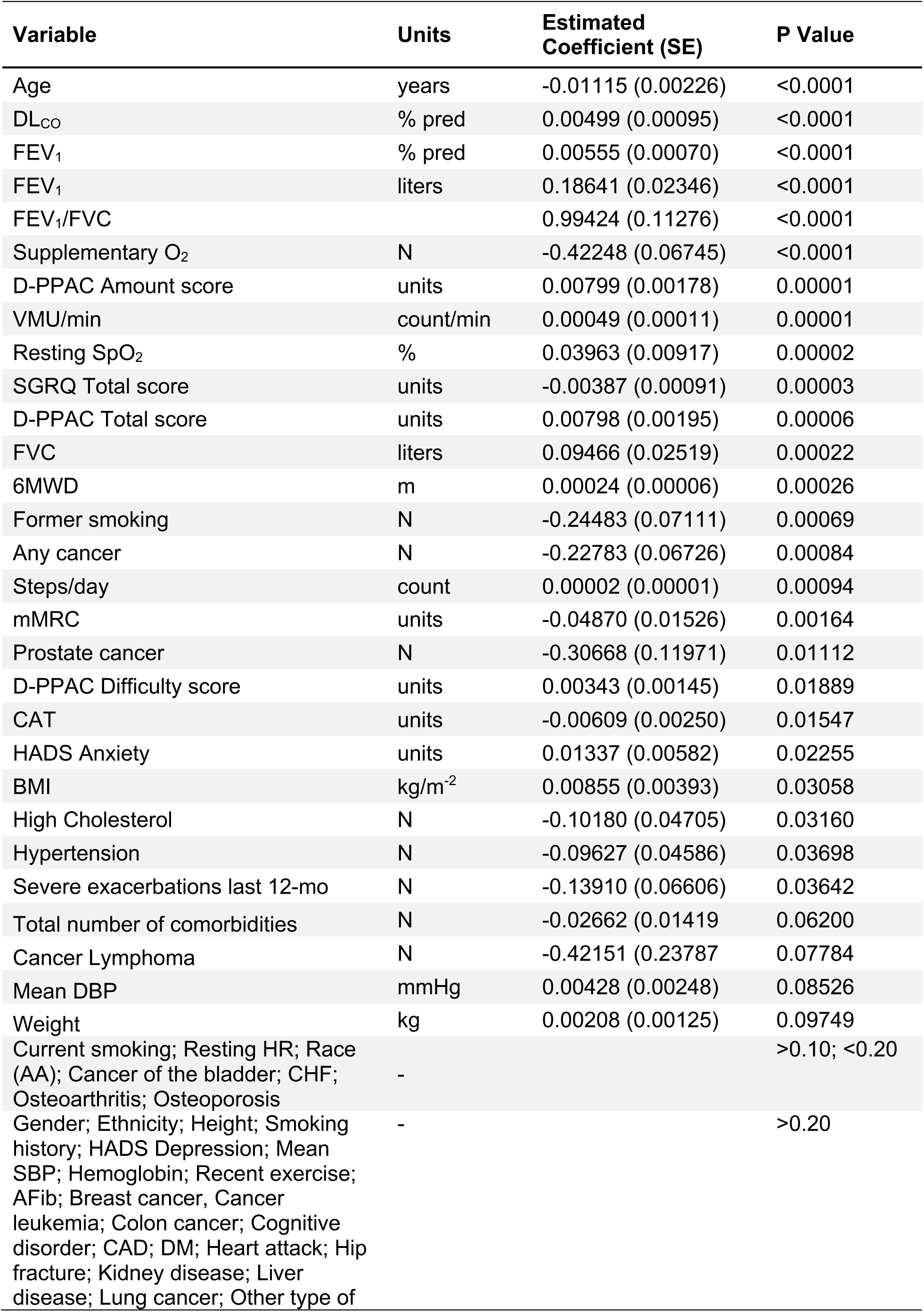
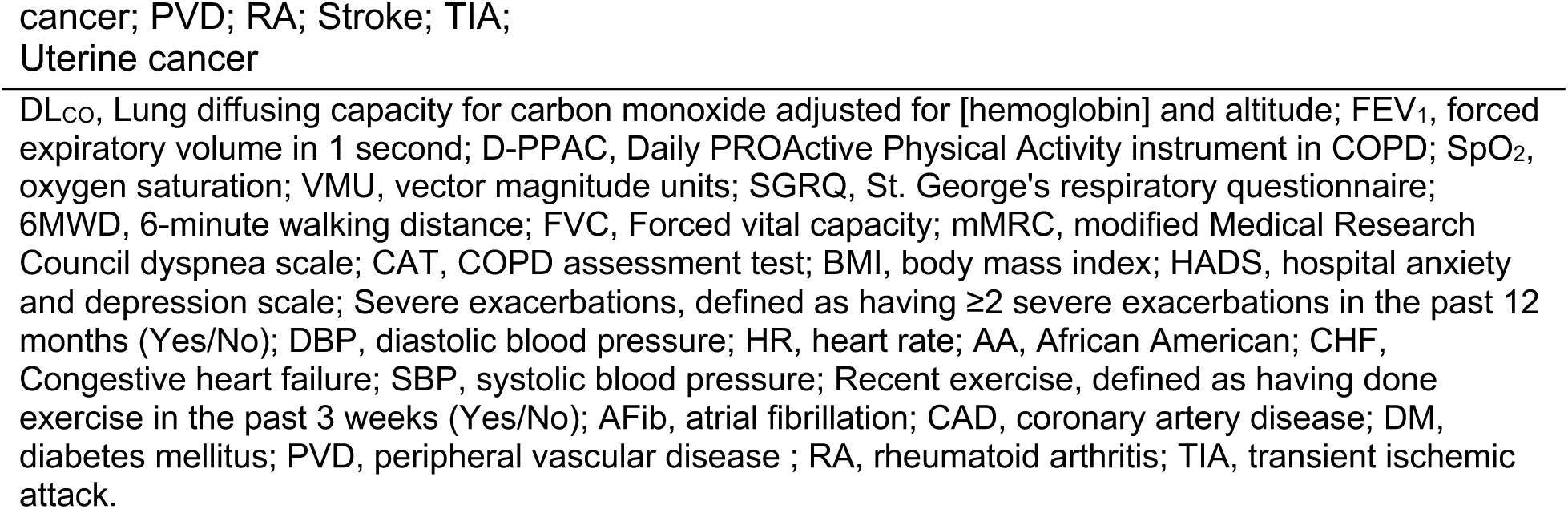
Univariate regression of association between muscle oxygen consumption recovery rate constant (*k*; proportional to muscle oxidative capacity; log transformed) and clinical and behavioral variables in the analysis cohort (n=241).

| Variable | Units | Estimated Coefficient (SE) | P Value |
| --- | --- | --- | --- |
| Age | years | -0.01115 (0.00226) | <0.0001 |
| DL<sub>CO</sub> | % pred | 0.00499 (0.00095) | <0.0001 |
| FEV<sub>1</sub> | % pred | 0.00555 (0.00070) | <0.0001 |
| FEV<sub>1</sub> | liters | 0.18641 (0.02346) | <0.0001 |
| FEV<sub>1</sub>/FVC |  | 0.99424 (0.11276) | <0.0001 |
| Supplementary O<sub>2</sub> | N | -0.42248 (0.06745) | <0.0001 |
| D-PPAC Amount score | units | 0.00799 (0.00178) | 0.00001 |
| VMU/min | count/min | 0.00049 (0.00011) | 0.00001 |
| Resting SpO<sub>2</sub> | % | 0.03963 (0.00917) | 0.00002 |
| SGRQ Total score | units | -0.00387 (0.00091) | 0.00003 |
| D-PPAC Total score | units | 0.00798 (0.00195) | 0.00006 |
| FVC | liters | 0.09466 (0.02519) | 0.00022 |
| 6MWD | m | 0.00024 (0.00006) | 0.00026 |
| Former smoking | N | -0.24483 (0.07111) | 0.00069 |
| Any cancer | N | -0.22783 (0.06726) | 0.00084 |
| Steps/day | count | 0.00002 (0.00001) | 0.00094 |
| mMRC | units | -0.04870 (0.01526) | 0.00164 |
| Prostate cancer | N | -0.30668 (0.11971) | 0.01112 |
| D-PPAC Difficulty score | units | 0.00343 (0.00145) | 0.01889 |
| CAT | units | -0.00609 (0.00250) | 0.01547 |
| HADS Anxiety | units | 0.01337 (0.00582) | 0.02255 |
| BMI | kg/m<sup>-2</sup> | 0.00855 (0.00393) | 0.03058 |
| High Cholesterol | N | -0.10180 (0.04705) | 0.03160 |
| Hypertension | N | -0.09627 (0.04586) | 0.03698 |
| Severe exacerbations last 12-mo | N | -0.13910 (0.06606) | 0.03642 |
| Total number of comorbidities | N | -0.02662 (0.01419) | 0.06200 |
| Cancer Lymphoma | N | -0.42151 (0.23787) | 0.07784 |
| Mean DBP | mmHg | 0.00428 (0.00248) | 0.08526 |
| Weight | kg | 0.00208 (0.00125) | 0.09749 |
| Current smoking; Resting HR; Race (AA); Cancer of the bladder; CHF; Osteoarthritis; Osteoporosis | - |  | >0.10; <0.20 |
| Gender; Ethnicity; Height; Smoking history; HADS Depression; Mean SBP; Hemoglobin; Recent exercise; AFib; Breast cancer, Cancer leukemia; Colon cancer; Cognitive disorder; CAD; DM; Heart attack; Hip fracture; Kidney disease; Liver disease; Lung cancer; Other type of | - |  | >0.20 |

DL<sub>CO</sub>, Lung diffusing capacity for carbon monoxide adjusted for [hemoglobin] and altitude; FEV<sub>1</sub>, forced expiratory volume in 1 second; D-PPAC, Daily PROActive Physical Activity instrument in COPD; SpO<sub>2</sub>, oxygen saturation; VMU, vector magnitude units; SGRQ, St. George's respiratory questionnaire; 6MWD, 6-minute walking distance; FVC, Forced vital capacity; mMRC, modified Medical Research Council dyspnea scale; CAT, COPD assessment test; BMI, body mass index; HADS, hospital anxiety and depression scale; Severe exacerbations, defined as having $\geq 2$ severe exacerbations in the past 12 months (Yes/No); DBP, diastolic blood pressure; HR, heart rate; AA, African American; CHF, Congestive heart failure; SBP, systolic blood pressure; Recent exercise, defined as having done exercise in the past 3 weeks (Yes/No); AFib, atrial fibrillation; CAD, coronary artery disease; DM, diabetes mellitus; PVD, peripheral vascular disease ; RA, rheumatoid arthritis; TIA, transient ischemic attack.

These 24 variables were then filtered by 3 machine learning algorithms to 11 variables with consistently highest importance (i.e., smallest cross-validated mean-squared error after machine learning filtering) on 161 (68.9% of completers) participants with complete assessments (‘analysis cohort’: 27 were missing primary outcome; 53 were missing predictor variables; Figure 1).

Multivariable regression predictors of *k* are shown in Table 3. FEV_1_%predicted, age and race were significant predictors of *k* (R^2^=0.26). Model coefficients equate a 10% lower FEV_1_%predicted to 4.4% lower *k*, or 10-years of aging to 9.7% lower *k*. Moreover, individuals of African American race had significantly 11% lower *k* than Non-Hispanic White peers with same smoking history. When radiography was included (‘CT-scan cohort’, n=118, 49.0% of study completers), FEV_1_%predicted and age remained the only significant predictors of *k* (R^2^=0.24).

**Table 3.** Multivariable regression clinical and behavioral predictors of muscle oxygen consumption recovery rate constant (*k*; proportional to muscle oxidative capacity) (n=161)

| Variable | Units | Estimated Coefficient | P Value |
| --- | --- | --- | --- |
| FEV<sub>1</sub> | % pred | 0.0045 | **0.0003** |
| Age | years | -0.0102 | **0.0005** |
| Race | AA | -0.1059 | **0.0472** |
| Resting SpO<sub>2</sub> | % | 0.0153 | 0.1299 |
| BMI | kg/m<sup>-2</sup> | 0.0049 | 0.2570 |
| DL<sub>CO</sub> | % pred | -0.001 | 0.4623 |
| Severe exacerbations last 12-months | N | 0.0294 | 0.6836 |
| Current smoking | N | -0.0353 | 0.6732 |
| VMU/min | count/min | <0.0001 | 0.7377 |
| Supplementary O<sub>2</sub> | N | -0.0327 | 0.7403 |
| D-PPAC total score | units | 0.0002 | 0.9302 |

FEV<sub>1</sub>, forced expiratory volume in 1 second; AA, African American race; SpO<sub>2</sub>, oxygen saturation; BMI, body mass index; DL<sub>CO</sub>, lung diffusing capacity for carbon monoxide adjusted for [hemoglobin] and altitude; Severe exacerbations, defined as having ≥2 severe exacerbations in the past 12 months (Yes/No); VMU, vector magnitude units; D-PPAC, Daily PROActive Physical Activity instrument in COPD.

To our surprise, none of our models retained any PA-related variable as a predictor of *k*. To corroborate this finding, we performed a sensitivity analysis on VMU/min (see details in Supplementary material). First, we removed each predictor in the model (Table 3) and found that VMU/min remained non-significant (p>0.222-0.984; Table S4). Second, we identified the predictors correlated with VMU/min (these were FEV_1_ %predicted, D-PPAC total score, supplementary O_2_ and current smoking) and investigated the significance of VMU/min when two of these correlated predictors were removed at the same time. Interestingly, VMU/min remained non-significant regardless of which two of the four correlated predictors were removed. The lowest p-value occurred when FEV_1_ %predicted and D-PPAC total were removed (p=0.100; adjusted R^2^=0.18), leaving age as the only significant predictor (p=0.010; Table S5). Similar results were found when the analysis was run using the CT-scan cohort (p=0.151; adjusted R^2^=0.19; Table S6). These analyses confirmed that PA is not a predictor of *k*.

## DISCUSSION

This study sought to identify predictors of locomotor muscle oxidative capacity in a large group of smokers with and without COPD and never smoker individuals. We hypothesized that low PA and age would be two of the strongest predictors of muscle oxidative impairment in people with COPD. We found that muscle oxidative capacity assessed by NIRS positively associated with lung function (FEV_1_%predicted), consistent with muscle biopsy studies[5, 6, 21], and negatively associated with age, consistent with some cross-sectional and longitudinal studies of aging[22-24]. However, our regression model demonstrated that free-living physical activities or COPD radiographic manifestations were not significant predictors of muscle oxidative capacity.

Muscle oxidative capacity is a key determinant of exercise capacity in health and COPD; it responds to endurance exercise training, contributing to improved exercise tolerance and symptom relief[25, 26]. Impaired mitochondrial oxidative capacity also associates with increased free radical production, systemic protein and DNA oxidation[27, 28], and reduced circulating di- or tri-acylglycerides in severe COPD[29]. In our completers cohort, we found that, compared to normal spirometry smokers, muscle oxidative capacity was 17.9% lower in mild COPD and 34-37% lower in severe COPD. Magnitude of these impairments are similar to previous reports in smaller cohorts[6, 30].

In absence of smoking or COPD, loss of muscle oxidative capacity is typically ascribed to aging and/or low PA. Some argue that muscle oxidative capacity loss may be protected by PA, such that models accounting for the lower PA associated with older age can explain much of muscle oxidative function loss[31, 32]. It is striking that several studies of older individuals do not find this association[24, 33, 34]. The Study of Muscle Mobility and Aging (SOMMA), which enrolled 697 community-dwelling elderly (≥70 years) with 4-m gait speed ≥0.6 m/s, showed significant association between accelerometer-derived PA and muscle oxidative capacity after age and sex adjustment, but not after accounting for other covariates including race, BMI and smoking status[24]. Additionally, in SOMMA increasing age was associated with reduced muscle oxidative capacity only in males and adjusting for PA had negligible impact on this association. Similarly, in our study including COPD patients, muscle oxidative impairment could not be explained by associated reductions in PA. Despite identifying VMU/min and D-PACC total score as significant correlates in univariate analyses, in multivariable analysis no activity variable was retained as a *k* predictor. This was further explored in a *post hoc* which consistently confirmed that PA was not a predictor of *k* in our study. Our finding of no association between PA and muscle oxidative capacity in COPD, is supported by a recent small study of nine COPD and activity-matched controls that found PA could not explain differences in muscle citrate synthase activity, mitochondrial respiratory function, or oxidative stress in quadriceps biopsies[35]. In fact, in that study, as in our 161 participants, spirometric variables were the major correlate of muscle oxidative impairment.

Another feature of our data supporting that low PA may not determine low muscle oxidative capacity in COPD, is that smokers with PRISm had significantly reduced activity behaviors (both VMU/min and steps/day were similar between PRISm and moderate and severe COPD), but muscle oxidative capacity in PRISm was not different from Controls (Figure 2a). Normal muscle oxidative capacity in PRISm, despite reduced PA, also counters an activity-based explanation for muscle oxidative impairment in COPD.

While cigarette smoke contains many mitotoxic components (e.g., carbon monoxide, cyanide) that may directly impair muscle mitochondrial function, we found no relation of muscle oxidative impairment to smoke exposure *per se* in normal spirometry participants (Figure 3a and 3b)[36-38]. This may be because exposure (≥1 cigarette within 2-hours) was too small, that activity of inhibited proteins of the mitochondrial respiratory chain (e.g., cytochrome c oxidase) are normally in excess and therefore reduced activity did not reduce overall oxidative capacity, or that typical smoke exposure levels do not directly affect skeletal muscle mitochondrial function. Nevertheless, smoke exposure *per se* could not explain muscle oxidative function impairments observed in this study.

African American race also associated with lower *k* (p=0.0472). To our knowledge, this is the largest study of muscle oxidative capacity in African Americans (n=110) and our results indicate that, among COPD patients, African Americans have 11% lower *k* than Non-Hispanic White adjusted for FEV_1_%predicted and age. Consistent with our findings, this difference in *k* could not be explained by low PA; neither steps/day (p=0.169) not VMU/min (p=0.136) differed between the two race groups.

FEV_1_%predicted, age and race were identified as muscle oxidative capacity predictors, but only explained ∼26% of the variance. Our model showed that FEV_1_%predicted is the strongest *k* predictor (p=0.0003; Table 3), and that 10% lower FEV_1_%predicted equated to 4.4% lower *k* i.e., about a fifth of the average decline between normal spirometry and severe COPD at FEV_1_ <50% predicted. This contradicts previous suggestions that lung function, in itself, is not a major factor responsible for lower limb muscle dysfunction in COPD[9]. While we did not address how pulmonary obstruction might explain locomotor muscle oxidative capacity impairment, we speculate that pulmonary and systemic inflammation in COPD may influence muscle mitochondrial biogenic and angiogenic pathways e.g., PGC-1a and VEGF expression, and mediate muscle mitochondrial loss, as has been demonstrated in smoke exposed mice[39]. Overall, our findings seem to support the myopathy theory[2, 30] in explaining etiology of skeletal muscle impairment in COPD.

Although our cohort was the among largest to evaluate contributors to muscle oxidative capacity in COPD, there was 33% missingness in the combination of muscle oxidative capacity and PA data that somewhat reduced statistical power. Future studies should focus on potential mediators of the connection between pulmonary obstruction and muscle oxidative capacity as modifiable variables to protect against muscle function loss. Also, our study design did not include assessment of muscle mass or strength. Given the strong association between muscle mass and mortality in COPD[40], identifying whether protecting against muscle oxidative function loss in COPD can protect against muscle atrophy is of considerable interest.

## Conclusion

Our data confirmed that older individuals with stable moderate to severe COPD have an 18-37% loss in locomotor skeletal muscle oxidative capacity, compared to smokers with normal spirometry, which could not be explained by lower physical activity. Pulmonary obstruction, older age and race were the strongest predictors of low muscle oxidative capacity in smokers with COPD.

## Supporting information

Supplementary Material_Adami et al.2025

## Data Availability

All data produced in the present study are available upon reasonable written request send to the corresponding author.

## ACKNOWLEDGEMENTS

We would like to thank all the participants for the time and dedication to the study, and the members of Respiratory Research Center at Harbor-UCLA for their support in coordinating the study enrolment and visits.

## ETHICAL GUIDELINES STATEMENT

Participants were informed about study procedures and risks before giving written informed consent and start the study protocol. The study was approved by the Institutional Review Board of The Lundquist Institute at Harbor-UCLA Medical Center (20403-01) and conducted in accordance with the ethical standards laid down in the 1964 Declaration of Helsinki and its later amendments.

