## Supplementary Material_Adami et al.2025 for "SEVERITY OF LUNG OBSTRUCTION AND OLDER AGE, BUT NOT PHYSICAL ACTIVITY, PREDICT LOCOMOTOR MUSCLE OXIDATIVE IMPAIRMENT IN COPD"

**This appendix has been provided by the authors to give readers additional information about their work.**

---

### METHODS

#### Participants

The main inclusion criteria for all study participants were: male and female, and aged 45 to 80 years old. Smokers were defined as individuals with ≥10 pack-years history of cigarette smoking. Never smokers were defined as individuals with a lifetime <100 cigarettes, <52 cigars, or <12oz. pipe tobacco smoked; and the absence of a physician diagnosed history of respiratory disease.

For participants enrolled in the COPDGene parent study, being of African American or Non-Hispanic White race was an additional main inclusion criterion, which was not applied to those recruited at the Harbor-UCLA Medical Center Pulmonary outpatient clinic.

The main exclusions for all study participants were: other concomitant respiratory disorder (such as, but not limited to, diffuse bronchiectasis, cystic fibrosis, or interstitial lung disease); lung surgery with removal of a lobe or more (including lung volume reduction and lung transplantation); lung cancer, known or suspected; bronchoscopic lung volume reduction; pregnancy or suspected pregnancy; uncontrolled cancer, as defined as ongoing radiation therapy, ongoing chemotherapy, narcotics for pain control, or known metastatic disease; history of radiation therapy to the chest (other than radiation for breast cancer); use of antibiotics and/or systemic corticosteroids (new prescription or increased dose) for a COPD exacerbation or any lung infection within the last month; inability to use albuterol; participation in pulmonary rehabilitation within 18 months; inability to perform a spirometry.

For full list of criteria see protocol available at www.copdgene.org.

**Study design**

As part of Phase 2 of the COPDGene[1] (NCT00608764) study protocol, participants completed a series of evaluations including (among others) spirometry, lung diffusing capacity for carbon monoxide ($DL_{CO}$), inspiratory and expiratory chest CT imaging, six-minute walking distance (6WMD) and questionnaires related to medical and smoking history, occupational and educational background, symptoms (COPD Assessment Test, CAT; modified Medical Research Council Dyspnea scale, mMRC), health-related quality of life (St. George's Respiratory Questionnaire, SGRQ; 36-Item Short Form Health Survey, SF-36), and anxiety and depression (Hospital Anxiety and Depression Scale, HADS).

The *Muscle Health Study* assessments were the NIRS-based muscle oxidative capacity test, resting pulse oximetry, objectively assessed free-living physical activity (PA) using 7-day triaxial accelerometry (for details on sensor technical characteristics and algorithms, see e.g.[2]), and the Daily-PROactive Physical Activity in COPD (D-PPAC) instrument[3].

Once arriving at the laboratory, participants first completed assessments for the COPDGene study protocol before completing assessments for the *Muscle Health Study*. To avoid potential technical or procedural bias in the results, the 19 participants recruited from Harbor-UCLA Medical Center Pulmonary outpatient clinic (i.e., not enrolled in COPDGene; Figure 1) were offered the same evaluations and questionnaires, following the same testing order, as those used in the COPDGene study. All participants were asked to self-report their race and ethnicity using an intake questionnaire.

**Pulmonary function and Imaging**

Spirometry was performed in accordance with American Thoracic Society guidelines[4] using a dual-beam Doppler ultrasound-based spirometer (EasyOne Pro, Ndd Medical, Zürich, Switzerland), before and after administration of two puffs of metered dose albuterol sulfate. The $FEV_1$ and FVC were measured from the greatest values obtained from up to eight maximum expiratory maneuvers, where the greatest two measurements were within 150 mL. Lung diffusing capacity for carbon monoxide ($DL_{CO}$) was measured in accordance with European Respiratory Society/American Thoracic Society standards for single-breath measurements of carbon monoxide uptake in the lungs[5], and adjusted for haemoglobin and altitude. Relative (%predicted) values were calculated using Global Lung Initiative reference equations[6].

Chest CT scans were performed in the supine position during a single full inspiration (200 mAs) and at end-tidal exhalation (50 mAs) with a 64-detector scanner[1]. Percentage of emphysema (%Emphysema) was defined as percentage of lung voxels with $\leq$-950 Hounsfield units (HU) attenuation on the inspiratory scan, and %gas trapping as the percentage of the lung volume with voxels $\leq$-856 HU on the expiratory scan. Airway wall thickening was assessed from the percentage of wall area percent (%Wall area) of $4^{th}$ and $5^{th}$ generation airways, and the square root of wall area of a 10-mm perimeter airway (Pi10)[7-9].

**Daily-PROactive Physical Activity instrument in COPD**

Participants were asked to reply to the Daily-PROactive Physical Activity instrument in COPD (D-PPAC version)[3], an instrument developed to evaluate the experience of patients with COPD with PA. A paper format of the instrument was used for this study, and it was provided to participant, together with a set of instructions, at the same time the accelerometer was given.

Participants were instructed to record their experiences daily on the same days they wore the accelerometer.

The D-PPAC is a self-administered 7-item questionnaire. Five items relate to the difficulty or limitation in completing daily activities (such as difficulty in getting dressed), and two items relate to the amount of PA done during the day (such as amount of walking). Each item is rated on a 0-4 points scale, and the analysis returns two sub-domain scores, the 'difficulty' and the 'amount' scores, the latter further combined with the daily steps and VMU count data derived from the accelerometer. The final scoring is converted into a Rassh score on a 0 (worst) to 100 (best) scale. The sum of the two sub-domains divided by two provides a total score, ranging from 0 (worse) to 100 (much better)[10]. The D-PPAC instrument is a reliable, valid, and responsive measure in diverse COPD patients[10], with good test-retest reliability in smokers with COPD (ICC ≥0.88)[3].

**Comorbidities**

Presence of comorbidities other than COPD were collected using the COPDGene Visit 2 Medical History Form (see copdgene.org; Phase 2 Study Documents). Participants were asked to report whether they had *"ever been told by a physician that they had*" the listed conditions.

The following conditions were considered: atrial fibrillation, breast cancer, cancer leukemia, cancer lymphoma, cancer of the bladder, cancer of the kidney, cancer of the throat and mouth, colon cancer, cognitive disorder, congestive heart failure, connective tissue disorder, coronary artery disease, diabetes mellitus, heart attack, high cholesterol, hip fracture, hypertension, kidney disease, liver disease, lung cancer, osteoarthritis, osteoporosis, other type of cancer, ovarian cancer, pancreatic cancer, peripheral vascular disease, prostate cancer, rheumatoid arthritis, stroke, transient ischemic attack (TIA), uterine cancer. Two additional variables were also calculated: 1) the total number of comorbidities for each participant; 2) the total count of participants within the completers cohort who reported at least one diagnosis of cancer among those considered (named "Any cancer"; see Table S1).

**Statistical Analyses**

Data are reported as mean(SD) for continuous variables, counts and percentage for discrete variables. One-way ANOVA with Dunnett's *post hoc*, Chi-square Test, Fisher's Exact Test of Independence were used to detect differences from smokers with normal spirometry (termed 'Controls'), as appropriate.

To determine association of predictor variables with *k*, after excluding variables with >30% missing data, univariate linear regression analysis was run with log-transformed *k* to satisfy linear regression assumptions. Then, all variables with $p \leq 0.20$ in univariate analysis were further filtered by machine learning-based feature selection methods such that more complex dependencies and interactions among predictors' effects on the response variable could be taken into consideration. Of the ten comorbidities with $p \leq 0.20$ in univariate analysis, seven (any cancer, cancer of the bladder, high cholesterol, hypertension, osteoarthritis, osteoporosis, total number of comorbidities) were collinear with PA-related variables (either VMU/min and/or steps/day; $p \leq 0.10$) and therefore were excluded from further analysis. The remaining three were excluded for low prevalence within the 'completers cohort' (cancer lymphoma n=2 (0.8%); congestive heart failure n=15 (6.2%); prostate cancer n=8 (3.3%)).

Three machine learning-based feature selection methods were employed. The first included the Least Absolute Shrinkage and Selection Operator (LASSO), a form of penalized regression incorporating an absolute value penalty term for regression coefficients in its objective function, with the goal to shrink certain coefficients to zero, thus facilitating variable selection within linear models[11]. The other two were tree-based algorithms, Random Forest[12], a method employing a bagging approach to constructing multiple decision trees in parallel, and the eXtreme Gradient Boosting (XGBoost), a method employing a boosting approach to sequentially building decision trees, each one aiming to correcting errors made by its predecessors[13]. For tree-based algorithms, a series of nodes within a tree are determined to split the feature space into multiple subspaces, and the predicted value for each subspace is calculated. The choice of the splitting variable for a node is typically based on quantitative criteria such as the improvement of node purity or the reduction in mean squared loss. In turn, the variable importance is conveniently measured as a variable's total contribution to the splitting of nodes across all trees according to the quantitative criterion. During the implementation of the three machine learning algorithms, their hyperparameters were selected based on the results of 5-fold cross-validation. The machine learning-based feature selection methods were run on the analysis cohort (n=161; 68.9% of 241 study completers). A *t*-test for unpaired groups showed no differences between the sub-group of 161 participants compared to the 241 who completed the study, for all variables investigated in this study.

By applying both the linear model LASSO and tree-based models Random Forest and XGBoost, we sought to measure the importance of each predictor of *k* in both a linear setting and a tree setting where interactions among predictors were naturally considered. A total of 13 predictors were identified and regarded as the candidate variables to be included in the multivariable linear regression model.

Before fitting the multivariable linear regression model, a variance inflation factor (VIF) for each predictor was computed. According to the screening results, two predictors with VIF values larger than 4 were removed from the model to address the collinearity issue. The multicollinearity of these two predictors ($FEV_1$/FVC and number of steps per day) was also confirmed by their extremely high pairwise correlation coefficients with retained predictors ($FEV_1$%predicted and VMU/min, respectively).

Finally, the remaining top 11 variables identified were entered into a multivariable linear regression to identify predictors of *k*. Due to a relatively large number of participants (43 individuals) who opted out of CT imaging, a second multivariable linear regression analysis was performed on participants with complete data including CT imaging (Figure 1).

After completing the primary analyses, in order to better understand the influence of significant predictors of *k*, we also explored the association of race (African American or Non-Hispanic White) with steps/day and with VMU/min within the analysis cohort (n=161). Steps/day and VMU/min were first log-transformed to satisfy linear regression assumptions, and then adjusted for $FEV_1$%predicted and age, respectively. In addition, we explored the sensitivity of the insignificance of VMU/min in the model. For this, we sequentially excluded each model predictor (Table 3) and investigated its effect on the coefficient and p-value estimation for VMU/min (results reported in Table S4). Next, we evaluated the correlation between VMU/min and each model predictor. For continuous predictors, we used Pearson correlation coefficient; for categorical predictors, we used 2-sample t-test or ANOVA. This analysis showed that VMU/min was moderately correlated with $FEV_1$ %predicted, D-PPAC total score, supplemental $O_2$ use and current smoking. The sensitivity of the insignificance of VMU/min was also evaluated by examining the effect on its p-value estimation when two of the four correlated predictors were excluded. By excluding $FEV_1$ %predicted and D-PPAC total score, VMU/min reached its lowest p-value of 0.100 in the predictive model ($R^2$=0.18; Table S5), although it remained non-significant. A similar result was found when the CT-scan cohort was used (p=0.151; $R^2$=0.19; Table S6).

The significance level (α) was set at 0.05. Analyses were performed using SPSS v28 (IBM, Chicago, IL, USA) and R (R 4.0.5); graphical representations with Prism (GraphPad, San Diego, CA, USA).

### SUPPLEMENTARY TABLES

**Table S1**. **Comorbidities in the cohort completing the study protocol (completers cohort; n=241)**. Comparison among smokers with normal spirometry (reference group; 'Controls'), never smokers, preserved ratio impaired spirometry, and patients with COPD (subdivided by disease severity). Conditions are listed in alphabetical order.

| | COHORT | Never Smokers | Normal Spirometry Smokers (Controls) | PRISm | COPD $FEV_1$ %predicted | | | |
|---|---|---|---|---|---|---|---|---|
| | | | | | > 80 | 50-79 | 30-49 | < 30 |
| N. | **241** | 26 | 76 | 21 | 30 | 40 | 35 | 13 |
| **Condition (N (%))** | | | | | | | | |
| Atrial fibrillation | **7 (2.9)** | 0 (0.0) | 0 (0.0) | 1 (5.0) | 0 (0.0) | 2 (5.0) | 3 (8.6) | 1 (7.7) |
| Any cancer | **28 (11.6)** | 1 (3.9) | 7 (9.2) | 1 (5.0) | 3 (10.0) | 4 (10.0) | 9 (25.7) | 3 (23.1) |
| Breast cancer | **4 (1.7)** | 0 (0.0) | 1 (1.3) | 0 (0.0) | 0 (0.0) | 1 (2.5) | 2 (5.7) | 0 (0.0) |
| Cancer leukemia | **1 (0.4)** | 0 (0.0) | 0 (0.0) | 0 (0.0) | 0 (0.0) | 1 (2.5) | 0 (0.0) | 0 (0.0) |
| Cancer lymphoma | **2 (0.8)** | 0 (0.0) | 0 (0.0) | 0 (0.0) | 1 (3.3) | 1 (2.5) | 0 (0.0) | 0 (0.0) |
| Cancer of the bladder | **2 (0.8)** | 0 (0.0) | 0 (0.0) | 0 (0.0) | 1 (3.3) | 0 (0.0) | 1 (2.9) | 0 (0.0) |
| Cancer of the throat and mouth | **0 (0.0)** | 0 (0.0) | 0 (0.0) | 0 (0.0) | 0 (0.0) | 0 (0.0) | 0 (0.0) | 0 (0.0) |
| Colon cancer | **3 (1.3)** | 0 (0.0) | 2 (2.6) | 0 (0.0) | 1 (3.3) | 0 (0.0) | 0 (0.0) | 0 (0.0) |
| Cognitive disorder | **3 (1.3)** | 1 (3.9) | 1 (1.3) | 0 (0.0) | 0 (0.0) | 0 (0.0) | 1 (2.9) | 0 (0.0) |
| Congestive heart failure | **15 (6.2)** | 0 (0.0) | 3 (4.0) | 3 (15.0) | 0 (0.0) | 5 (12.5) | 3 (8.6) | 1 (7.7) |
| Connective tissue disorder | **0 (0.0)** | 0 (0.0) | 0 (0.0) | 0 (0.0) | 0 (0.0) | 0 (0.0) | 0 (0.0) | 0 (0.0) |
| Coronary artery disease | **11 (4.6)** | 1 (3.9) | 4 (5.3) | 1 (5.0) | 1 (3.3) | 0 (0.0) | 3 (8.6) | 1 (7.7) |
| Diabetes mellitus | **32 (13.3)** | 1 (3.9) | 12 (15.8) | 6 (30.0) | 0 (0.0) | 5 (12.5) | 7 (20.0) | 1 (7.7) |
| Heart attack | **13 (5.4)** | 0 (0.0) | 5 (6.6) | 2 (10.0) | 1 (3.3) | 2 (5.0) | 2 (5.7) | 1 (7.7) |
| High cholesterol | **87 (36.1)** | 7 (26.9) | 24 (31.6) | 6 (30.0) | 11 (36.7) | 20 (50.0) | 14 (40.0) | 5 (38.5) |
| Hip fracture | **8 (3.3)** | 0 (0.0) | 3 (4.0) | 0 (0.0) | 1 (3.3) | 0 (0.0) | 2 (5.7) | 2 (15.4) |
| Hypertension | **121 (50.2)** | 8 (30.8) | 36 (47.4) | 14 (70.0) | 11 (36.7) | 24 (60.0) | 22 (62.9) | 6 (46.2) |

| | | | | | | | | |
|---|---|---|---|---|---|---|---|---|
| Kidney cancer | **0 (0.0)** | 0 (0.0) | 0 (0.0) | 0 (0.0) | 0 (0.0) | 0 (0.0) | 0 (0.0) | 0 (0.0) |
| Kidney disease | **4 (1.7)** | 1 (3.9) | 0 (0.0) | 2 (10.0) | 0 (0.0) | 1 (2.5) | 0 (0.0) | 0 (0.0) |
| Liver disease | **9 (3.7)** | 0 (0.0) | 7 (9.2) | 0 (0.0) | 1 (3.3) | 0 (0.0) | 0 (0.0) | 1 (7.7) |
| Lung cancer | **2 (0.8)** | 0 (0.0) | 0 (0.0) | 0 (0.0) | 0 (0.0) | 0 (0.0) | 1 (2.9) | 1 (7.7) |
| Osteoarthritis | **37 (15.4)** | 2 (7.7) | 12 (15.8) | 3 (15.0) | 5 (16.7) | 6 (15.0) | 7 (20.0) | 2 (15.4) |
| Osteoporosis | **21 (8.7)** | 1 (3.9) | 4 (5.3) | 1 (5.0) | 2 (6.7) | 2 (5.0) | 6 (17.1) | 5 (38.5) |
| Other type of cancer | **2 (0.8)** | 0 (0.0) | 1 (1.3) | 0 (0.0) | 0 (0.0) | 0 (0.0) | 1 (2.9) | 0 (0.0) |
| Ovarian cancer | **1 (0.4)** | 0 (0.0) | 0 (0.0) | 1 (5.0) | 0 (0.0) | 0 (0.0) | 0 (0.0) | 0 (0.0) |
| Pancreatic cancer | **0 (0.0)** | 0 (0.0) | 0 (0.0) | 0 (0.0) | 0 (0.0) | 0 (0.0) | 0 (0.0) | 0 (0.0) |
| Peripheral vascular disease | **2 (0.8)** | 0 (0.0) | 0 (0.0) | 0 (0.0) | 1 (3.3) | 0 (0.0) | 0 (0.0) | 1 (7.7) |
| Prostate cancer | **8 (3.3)** | 1 (3.9) | 2 (2.6) | 0 (0.0) | 0 (0.0) | 1 (2.5) | 4 (11.4) | 0 (0.0) |
| Rheumatoid arthritis | **17 (7.1)** | 1 (3.9) | 5 (6.6) | 3 (15.0) | 3 (10.0) | 3 (7.5) | 1 (2.9) | 1 (7.7) |
| Stroke | **3 (1.3)** | 0 (0.0) | 2 (2.6) | 0 (0.0) | 0 (0.0) | 1 (2.5) | 0 (0.0) | 0 (0.0) |
| Transient ischemic attack | **5 (2.1)** | 2 (7.7) | 1 (1.3) | 1 (5.0) | 0 (0.0) | 1 (2.5) | 0 (0.0) | 0 (0.0) |
| Uterine cancer | **3 (1.3)** | 0 (0.0) | 1 (1.3) | 0 (0.0) | 0 (0.0) | 0 (0.0) | 0 (0.0) | 2 (15.4) |

Data are N (%). $FEV_1$, forced expiratory volume in 1 second; PRISm, preserved ratio impaired spirometry. Spirometric variables are postbronchodilator.

**Table S2. Symptoms and quality of life related questionnaires of the cohort completing the study protocol (completers cohort; n=241).** Comparison among smokers with normal spirometry (reference group; ‘Controls’), never smokers, preserved ratio impaired spirometry, and patients with COPD (subdivided by disease severity).

| | COHORT | Never Smokers | Normal Spirometry Smokers (Controls) | PRISm | COPD $FEV_1$ %pred | | | |
|---|---|---|---|---|---|---|---|---|
| | | | | | > 80 | 50-79 | 30-49 | < 30 |
| N | **241** | 26 | 76 | 21 | 30 | 40 | 35 | 13 |
| mMRC | **1.2 (1.5)** | 0.5 (1.0) | 0.8 (1.3) | 1.3 (1.4) | 0.5 (1.0) | 1.5 (1.5) * | 2.8 (1.2) * | 3.0 (1.4) * |
| CAT | **12.9 (9.0)** | 4.5 (6.0) | 12.1 (8.0) | 13.1 (10.0) | 9.1 (5.8) | 15.1 (8.5) | 18.8 (8.6) * | 20.5 (9.0) * |
| SGRQ Total Score | **24.1 (24.2)** | 6.5 (14.8) | 14.0 (17.1) | 22.2 (22.2) | 13.8 (18.6) | 31.1 (21.9) * | 48.0 (20.2) * | 58.5 (19.6) * |
| Symptoms score | **24.7 (27.0)** | 5.9 (14.7) | 14.2 (22.1) | 24.0 (29.5) | 15.2 (22.6) | 37.7 (24.1) * | 46.5 (25.5) * | 48.8 (21.0) * |
| Active score | **35.3 (32.0)** | 10.5 (21.8) | 23.8 (25.6) | 37.0 (29.3) | 20.3 (23.5) | 43.9 (29.6) * | 64.6 (25.1) * | 79.3 (19.3) * |
| Impact score | **17.3 (21.8)** | 4.4 (11.4) | 8.2 (13.3) | 13.2 (10.1) | 9.6 (17.5) | 21.9 (20.3) * | 38.7 (21.1) * | 49.5 (24.2) * |
| SF-36 PF score | **64.4 (30.6)** | 85.4 (22.8) | 76.1 (23.5) | 58.8 (25.6) * | 73.0 (23.2) | 61.8 (30.9) * | 39.1 (26.6) * | 19.6 (23.7) * |
| RP score | **71.6 (30.7)** | 87.7 (22.0) | 81.7 (24.5) | 65.8 (27.7) | 81.7 (27.2) | 71.9 (28.8) | 48.2 (27.3) * | 27.9 (31.1) * |
| PCS score | **54.3 (11.2)** | 52.2 (8.0) | 48.6 (8.9) | 42.3 (9.4) * | 47.7 (9.9) | 44.0 (11.8) | 35.3 (9.1) * | 27.1 (8.8) * |
| MCS score | **50.1 (11.1)** | 53.2 (12.0) | 49.4 (10.2) | 50.4 (14.1) | 51.9 (10.5) | 50.8 (8.8) | 46.9 (9.6) | 44.7 (18.9) |
| HADS Anxiety | **5.2 (3.9)** | 4.8 (4.6) | 6.0 (3.8) | 5.5 (4.5) | 4.5 (3.7) | 4.1 (3.2) | 5.3 (3.7) | 5.9 (5.1) |
| HADS Depression | **4.4 (3.7)** | 3.7 (4.3) | 4.6 (3.4) | 4.1 (3.9) | 3.5 (3.5) | 3.9 (3.4) | 4.9 (3.7) | 6.8 (3.5) |

Data are mean (SD) if not otherwise specified. PRISm, preserved ratio impaired spirometry; mMRC, modified medical research council dyspnea scale; CAT, COPD assessment test; SGRQ, St. George's respiratory questionnaire; SF-36, Short-form 36-item questionnaire; PF, SF-36 subscale physical functioning; RP, SF-36 subscale role limitations due to physical health; PCS, SF-36 aggregate physical component score; MCS, SF-36 aggregate mental component score; HADS, hospital anxiety and depression scale. * p<0.05 versus normal spirometry smokers.

**Table S3**. **Pulmonary anatomic variables in the chest CT scan cohort (n=118)**. Comparison among smokers with normal spirometry (reference group; 'Controls'), never smokers, preserved ratio impaired spirometry, and patients with COPD (subdivided by disease severity).

| | COHORT | Never Smokers | Normal Spirometry Smokers (Controls) | PRISm | COPD $FEV_1$ %pred | | | |
|---|---|---|---|---|---|---|---|---|
| | | | | | > 80 | 50-79 | 30-49 | < 30 |
| N | **118** | 16 | 68 | 16 | 21 | 33 | 17 | 4 |
| %Emphysema | **17.3 (13.4)** | 7.1 (7.2) | 10.7 (7.2) | 8.8 (6.2) | 20.9 (11.8) * | 20.9 (12.6) * | 38.8 (8.9) * | 42.3 (7.2) * |
| %Gas Trapping | **20.5 (18.8)** | 10.8 (9.8) | 10.0 (8.6) | 8.0 (7.6) | 22.5 (12.1) * | 25.9 (16.1) * | 52.6 (14.4) * | 65.5 (7.0) * |
| %Wall Area | **48.7 (8.0)** | 44.5 (6.2) | 45.1 (7.3) | 55.3 (8.6) * | 48.5 (7.1) | 52.6 (6.7) * | 52.8 (7.2) * | 55.1 (6.7) * |
| Pi10 (mm) | **2.3 (0.5)** | 2.0 (0.3) | 2.0 (0.4) | 2.8 (0.5) * | 2.2 (0.4) | 2.6 (0.5) * | 2.7 (0.5) * | 2.9 (0.5) * |

Data are mean (SD). PRISm, preserved ratio impaired spirometry; $FEV_1$, forced expiratory volume in 1 second; Pi10, average wall thickness for a hypothetical airway of 10-mm lumen perimeter on CT. * $p<0.05$ versus normal spirometry smokers.

**Table S4. Sensitivity analysis for VMU/min.** Each variable included in the multivariate regression model (Table 3) was removed one at a time.

VMU/min remains non-significant in the regression no matter which single variable is excluded (analysis cohort; n=161).

| **Variable Excluded** | **VMU/min** | |
|---|---|---|
| | Estimated Coefficient | p-value |
| $FEV_1$ | 0.00011 | 0.3857 |
| Age | 0.00015 | 0.2220 |
| Race | 0.00005 | 0.7075 |
| Resting $SpO_2$ | 0.00005 | 0.6864 |
| BMI | <0.00001 | 0.9841 |
| $DL_{CO}$ | 0.00005 | 0.6561 |
| Severe exacerbations last 12-months | 0.00006 | 0.6447 |
| Current smoking | 0.00004 | 0.7629 |
| Supplementary $O_2$ | 0.00004 | 0.7224 |
| D-PPAC total score | 0.00009 | 0.4242 |

$FEV_1$, forced expiratory volume in 1 second; AA, African American race; $SpO_2$, oxygen saturation; BMI, body mass index; $DL_{CO}$, lung diffusing capacity for carbon monoxide adjusted for [hemoglobin] and altitude; Severe exacerbations, defined as having "≥" 2 severe exacerbations in the past 12 months (Yes/No); VMU, vector magnitude units; D-PPAC, Daily PROActive Physical Activity instrument in COPD

**Table S5** Multivariable regression clinical and behavioral predictors of muscle oxygen consumption recovery rate constant (*k*; proportional to muscle oxidative capacity) when $FEV_1$ %predicted and D-PPAC were removed after sensitivity analysis reported being correlates of VMU/min (n=161).

| **Variable** | **Units** | **Estimated Coefficient** | **P Value** |
|---|---|---|---|
| Age | years | -0.0071 | **0.0098** |
| Supplementary $O_2$ | N | -0.1581 | 0.0908 |
| VMU/min | count/min | 0.0002 | 0.0996 |
| Resting $SpO_2$ | % | 0.0142 | 0.1664 |
| Race | AA | -0.0636 | 0.1967 |
| BMI | $kg/m^{-2}$ | 0.0047 | 0.2609 |
| Current smoking | N | -0.0809 | 0.2721 |
| $DL_{CO}$ | % pred | 0.0009 | 0.4737 |
| Severe exacerbations last 12-months | N | -0.0234 | 0.7257 |

AA, African American race; VMU, vector magnitude units; $SpO_2$, oxygen saturation; BMI, body mass index; $DL_{CO}$, lung diffusing capacity for carbon monoxide adjusted for [hemoglobin] and altitude; Severe exacerbations, defined as having ≥2 severe exacerbations in the past 12 months (Yes/No).

**Table S6.** Multivariable regression clinical and behavioral predictors of muscle oxygen consumption recovery rate constant (*k*; proportional to muscle oxidative capacity) on chest CT scan cohort (n=118). $FEV_1$ %predicted and D-PPAC were removed after sensitivity analysis reported being correlates of VMU/min.

| **Variable** | **Units** | **Estimated Coefficient** | **P Value** |
|---|---|---|---|
| Age | years | -0.0094 | **0.0044** |
| Pi10 | mm | -0.1072 | 0.0675 |
| VMU/min | count/min | 0.0002 | 0.1512 |
| Supplementary $O_2$ | N | -0.1611 | 0.2043 |
| Race | AA | -0.0626 | 0.3309 |
| $DL_{CO}$ | % pred | -0.0002 | 0.9208 |
| Current smoking | N | -0.0682 | 0.4625 |
| Gas trapping | % | -0.0021 | 0.4666 |
| Resting $SpO_2$ | % | 0.0120 | 0.4722 |
| BMI | $kg/m^{-2}$ | 0.0031 | 0.5731 |
| Severe exacerbations last 12-months | N | -0.0046 | 0.9555 |

Pi10, average wall thickness for a hypothetical airway of 10-mm lumen perimeter on CT; VMU, vector magnitude units; AA, African American race; $DL_{CO}$, lung diffusing capacity for carbon monoxide adjusted for [hemoglobin] and altitude; $SpO_2$, oxygen saturation; BMI, body mass index; Severe exacerbations, defined as having ≥2 severe exacerbations in the past 12 months (Yes/No).

8. Patel BD, Coxson HO, Pillai SG, Agusti AG, Calverley PM, Donner CF, et al. Airway wall thickening and emphysema show independent familial aggregation in chronic obstructive

pulmonary disease. Am J Respir Crit Care Med. 2008;178:500-5. doi:10.1164/rccm.200801-059OC
